# Self-reported acceptance of a wearable activity monitor in persons with stroke

**DOI:** 10.1101/2024.12.14.24318939

**Authors:** Jamie Nam, Grace C Bellinger, Junyao Li, Margaret A French, Ryan T Roemmich

**Affiliations:** Department of Physical Medicine and Rehabilitation, Johns Hopkins University School of Medicine, Baltimore, MD 21205; Northwestern University Feinberg School of Medicine, Chicago, IL 60611; Department of Physical Therapy and Athletic Training, University of Utah, Salt Lake City, UT 84108; Center for Movement Studies, Kennedy Krieger Institute, Baltimore, MD 21205

**Author notes:** Contact information: Room 240 Kennedy Krieger Institute 716 North Broadway Baltimore, MD 21205. corresponding author Twitter: @GraceCBellinger (GCB), @MaggieFrenchDPT (MAF), @RyanRoemmich (RTR).

**Keywords:** stroke, wearables, mobility, technology acceptance, physical activity, remote monitoring, rehabilitation

## Abstract

**Background:** Wearable activity monitors offer clinicians and researchers accessible, scalable, and cost-effective tools for continuous remote monitoring of functional status. These technologies can complement traditional clinical outcome measures by providing detailed, minute-by-minute remotely collected data on a wide array of biometrics that include, as examples, physical activity and heart rate. There is significant potential for the use of these devices in rehabilitation after stroke if individuals will wear and use the devices; however, the acceptance of these devices by persons with stroke is not well understood.

**Objective:** In this study, we investigated the participant-reported acceptance of a commercially available, wrist-worn wearable activity monitor (the Fitbit Inspire 2) for remote monitoring of physical activity and heart rate in persons with stroke. We also assessed relationships between reported acceptance and adherence to wearing the device.

**Methods:** Sixty-five participants with stroke wore a Fitbit Inspire 2 for three months, at which point we assessed the acceptance of wearing the device using the Technology Acceptance Questionnaire (TAQ) inclusive of its seven dimensions (Perceived Usefulness (PU), Perceived Ease of Use (PEOU), Equipment Characteristics (EC), Privacy Concern (PC), Perceived Risk (PR), Facilitating Conditions (FC), and Subjective Norm (SN). We then performed Spearman’s correlations to assess relationships between acceptance and adherence to device wear, which we calculated as both the percentage of daily wear time and percentage of valid days the device was worn during the three weeks preceding TAQ administration.

**Results:** Most participants reported generally agreeable responses with high overall total TAQ scores across all seven dimensions that indicated strong acceptance of the device; “Agree” was the median response to 29 of the 31 TAQ statements. Participants generally found the device beneficial for their health, efficient for monitoring, easy to use and don/doff, and unintrusive to daily life. However, participant responses on the TAQ did not show significant positive correlations with measures of actual device wear time (all p>0.05).

**Conclusions:** This study demonstrates generally high self-reported acceptance of the Fitbit Inspire 2 among persons with stroke. Participants reported general agreement across all seven TAQ dimensions with minimal concerns interpreted as being directly relatable to post-stroke motor impairment (e.g., donning and doffing devices, using the device independently). However, the high self-reported acceptance scores did not correlate positively with measures of real-world device wear. Accordingly, it should not be assumed that persons with stroke will adhere to wearing these devices simply because they report high acceptability.

## Introduction

### Background

Wearable devices have the potential to advance how clinicians and researchers measure functional status by offering accessible, scalable, and cost-effective remote monitoring tools.[1,2] Traditional outcome measures provide only a discrete snapshot of an individual’s functional status;[3–5] emerging wearable technologies have the potential to address this issue by providing minute-by-minute, longitudinal data on physical activity, heart rate, and many other biometrics from daily life that extend beyond the clinical or laboratory setting. The use of wearable devices for remote monitoring could offer additional observational data on functional status measured directly in an individual’s daily environment.

Notably, wearable devices could provide valuable insight into recovery following neurologic damage (e.g., after stroke) by enabling granular, longitudinal assessment of activity, one of the components of the World Health Organization International Classification of Functioning, Disability, and Health model.[6] Approximately 80% of persons with stroke experience some form of motor impairment, and around 50% continue to have significant functional limitations six months post-stroke.[7–9] These limitations often translate into reduced daily activity, as persons with stroke generally walk approximately half as many steps each day as individuals without stroke.[10] Wearable devices provide an opportunity to monitor these functional changes remotely with insights on daily physical activity. However, the ability to perform remote monitoring after stroke using wearable devices is dependent on how well persons with stroke accept these devices.

### Prior Work

Wearable activity monitors like Fitbit devices generally show high acceptance among healthy individuals[11] and a range of patient groups, including older adults with cognitive and motor impairment.[12,13] For older adults with cognitive impairment, Fitbit-based interventions may improve motivation for physical activity and sleep, but success is dependent on interfaces that are easy to use,[14] reduce cognitive load,[15] and are beneficial to the user.[14] In studies of persons with minimal motor impairment, the lack of sustained Fitbit usage and challenges to acceptance have largely been attributed to technical issues – difficulties with empty batteries, broken devices, or lost devices–[16] rather than usability concerns. For example, users with multiple sclerosis reported frustration when syncing data between devices.[17]

To measure acceptance of wearable technologies across diverse clinical populations, researchers have extensively used the Technology Acceptance Model (TAM) due to its simplicity and strong empirical support[18,19], with the Technology Acceptance Questionnaire (TAQ)[13] serving as an extension of this framework with additional factors related to user acceptance. Numerous studies have validated this framework across many contexts, demonstrating its predictive power in understanding technology adoption behaviors.[18–22] While wearable activity monitors have demonstrated varying levels of acceptance across the general population, their perceived usability and effectiveness in individuals with stroke remain less explored, prompting our use of the TAQ to assess their suitability in this specific patient group.[23]

### Goal of This Study

Here, we aimed to assess the acceptance of wearable devices in persons with stroke. We also examined the relationship between acceptance and adherence to wearing the Fitbit device. We studied Fitbit devices specifically because these have been used extensively for remote monitoring of step count, heart rate, and energy expenditure – among other metrics – in many populations.[24] We hypothesized that 1) acceptance of the Fitbit devices would be variable across persons with stroke but would generally indicate that these technologies are acceptable, useful, and easy to use, and 2) acceptance would be significantly associated with real-world adherence to wearing the Fitbit device.

## Methods

### Recruitment

We recruited persons with stroke from the outpatient rehabilitation clinics at Johns Hopkins Hospital through clinician referrals through MyChart messages. The inclusion criteria for this study were: age 18 years and older; history of stroke (as confirmed by ICD-10 codes); ownership of a smartphone and in-home Wi-Fi to access the internet; walking as a primary form of mobility (with assistive devices allowed); and ability and willingness to install the Fitbit mobile application on their smartphone device. All participants provided oral consent, as approved by the Institutional Review Board at the Johns Hopkins University School of Medicine (IRB00247292).

After obtaining consent, the study team mailed a Fitbit Inspire 2 (Fitbit Inc., San Francisco, CA, USA) to participants and asked them to wear it for one year as part of a larger study.[25] This study focuses on a sub-analysis of the first three months of device wear, during which the Technology Acceptance Questionnaire (TAQ) was administered to assess participants’ experiences and perceptions of the device at the three-month time point. As an incentive, we allowed participants to keep the Fitbit following completion of the study.

### Study Instructions

We instructed participants to wear the Fitbit on their non-paretic (i.e., less impaired) wrist throughout the entire day, including during sleep; if participants had difficulty donning the device and lacked available assistance, we permitted them to wear it on the paretic wrist. We documented the paretic side and the wrist on which the Fitbit was worn. Then, we guided the participants over the phone to set up the device and install the Fitbit application on their smartphone. We instructed participants to remove the device only when showering or charging it. We also asked them to synchronize the device at least once per day using the Fitbit smartphone application. Data from the Fitbit were extracted by a custom-built application that is described elsewhere.[25,26] The study team incorporated notifications and reminders to assist with adherence to wearing the Fitbit, as detailed in our previous work.[25]

After a participant was enrolled for three months, the study team attempted to contact the participant up to three times to administer the TAQ, our metric of acceptance. Each contact attempt was documented in REDCap, which included the date and time of the call, the outcome of the attempt (e.g., reached, voicemail, no answer), the participant’s response to the assessment, and any notes or follow-up actions required. If the participant was reached, the team member administered the questionnaire verbally. However, if the participant was not reached after three attempts, no further attempts were made to administer the TAQ.

### Technology Acceptance Questionnaire (TAQ)

We used the TAQ – which includes dimensions for Perceived Usefulness (PU), Perceived Ease of Use (PEOU), Equipment Characteristics (EC), Privacy Concern (PC), Perceived Risk (PR), Facilitating Conditions (FC), and Subjective Norm (SN) – as established by Puri et al.[13] The full questionnaire is included as Multimedia Appendix 1 in Puri et al.[13] The TAQ consisted of 31 statements rated on a 5-point Likert scale where participants indicated their levels of agreement or disagreement with each statement. These 31 statements covered dimensions of perceived usefulness (PU; n=5 statements), perceived ease of use (PEOU, n=7 statements), facilitating conditions (FC; n=2 statements), subjective norm (SN; n=3 statements), equipment characteristics (EC; n=8 statements), privacy concern (PC; n=3 statements), and perceived risk (PR; n=3 statements). We summed the scores within each dimension (“Strongly Disagree” = 1 point, “Disagree” = 2 points, “Neutral” = 3 points, “Agree” = 4 points, “Strongly Agree” = 5 points). We note that, to ensure the reliability and validity of the measure, certain statements in the TAQ were negatively framed to mitigate acquiescence bias.[27] As is standard, we minimized possible response biases where higher numerical values represent lower agreement by applying a reverse coding procedure to these statements: each affected score was standardized by subtracting it from 6 for statements 2, 10, 17, 25, and 27. This was done prior to summing the scores within each dimension. We then calculated the total TAQ score by summing the scores from all items across all dimensions. In addition to the 31 statements, there are also six multiple choice questions addressing various aspects of device use that are not assigned to any domain.

### Fitbit Adherence

To assess adherence, we analyzed Fitbit data from the 21 days immediately preceding completion of the TAQ. This timeframe was selected because the TAQ items specifically reference user perceptions of their Fitbit over the prior three weeks. We identified Fitbit wear minutes as any minute with a non-zero heart rate through the accelerometry package[28] in R.[29] We calculated two primary adherence metrics. First, we calculated the percentage of average daily wear time as the percentage of time the device was worn each day by dividing the number of wear minutes by the total maximum amount of minutes in a day (1440) and multiplied by 100. These daily percentages were then averaged across the 21-day period. Second, we calculated percentage of valid wear days the number of days within the three-week window with at least 75% of wear time (i.e., 1080 or more minutes). The number of valid days was then divided by 21 (the total days in the assessment window) and multiplied by 100 to determine the percentage of valid days.

### Statistical Analysis

We calculated descriptive statistics for the 37 individual items of the TAQ (inclusive of the 31 Likert scale statements and 6 additional multiple-choice questions), the summed scores on each of the seven dimensions from the TAQ, and the summed total score of the TAQ. We also report descriptive statistics for the metrics of adherence. To assess relationships between Fitbit adherence and TAQ responses, we used Spearman’s correlations (due to the ordinal nature of the TAQ responses). We performed all statistical analyses using R version 4.4.1[29] with α=0.05.

## RESULTS

### Participants

Of the 108 persons with stroke that we contacted, 98 participants enrolled in the study (the remaining 10 individuals either did not meet the inclusion criteria or declined to participate). Among the enrolled participants, four were lost to follow-up (i.e., they could not be reached despite repeated attempts during the three-month study period) and eight withdrew voluntarily (i.e., they chose to discontinue their involvement after enrolling).

Furthermore, 21 participants did not complete the TAQ three months post-enrollment. As a result, we included 65 participants with stroke in the final analysis. We include a CONSORT diagram in Figure 1 and summarized study and excluded participant characteristics in Table 1.

**Figure 1.**
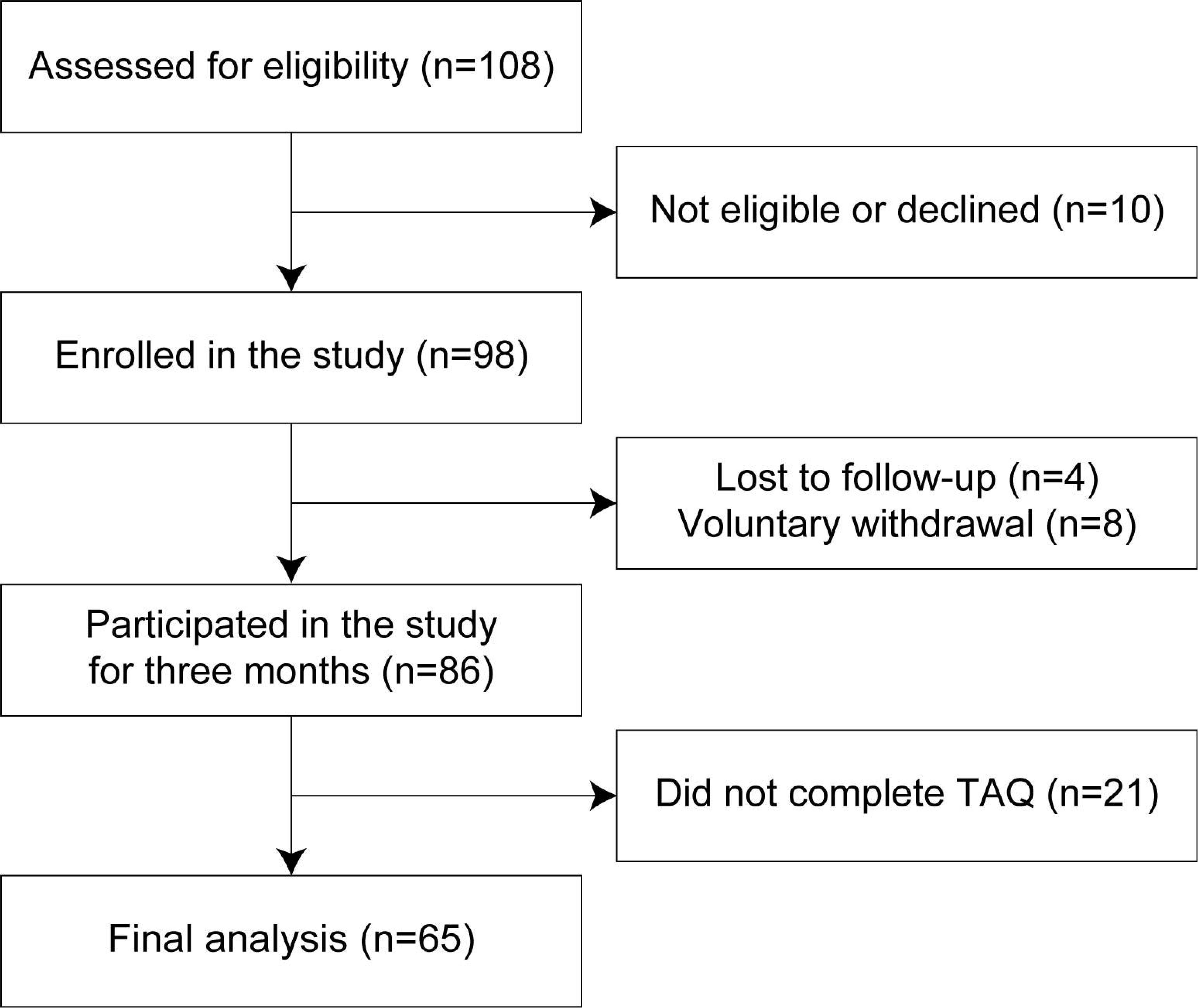
CONSORT diagram showing the enrollment and participation of participants at various stages throughout the study.

**Table 1.** Study participant characteristics.

|  |  | *Included (n=65)* | *Excluded (n=35)* |
| --- | --- | --- | --- |
|  |  | **Mean±SD** | **Mean±SD** |
| Age (years) |  | 62.4±12.4 | 60.5±12.5 |
| Days post-stroke at date of enrollment |  | 1053.9±1664.1 | 230.1±367.0 |
| Days between date of enrollment and date of TAQ completion |  | 127.8±42.9 | N/A |
|  |  | **N (%)** | **N (%)** |
| Sex |  |  |  |
|  | *Male* | 37 (56.9%) | 20 (57.1%) |
|  | *Female* | 28 (43.1%) | 15 (42.9%) |
| Race |  |  |  |
|  | *American Indian or Alaska Native* | 2 (3.1%) | 0 (0.0%) |
|  | *Asian* | 3 (4.6%) | 1 (2.9%) |
|  | *Black or African American* | 21 (32.3%) | 22 (62.9%) |
|  | *White/Caucasian* | 38 (58.5%) | 10 (28.6%) |
|  | *Multiple* | 1 (1.5%) | 2 (5.7%) |
| Ethnicity |  |  |  |
|  | *Hispanic or Latino* | 4 (6.2%) | 1 (2.9%) |
|  | *Not Hispanic or Latino* | 61 (93.8%) | 34 (97.1%) |
| Fitbit Wrist Worn* |  |  |  |
|  | *Paretic* | 15 (23.1%) | N/A |
|  | *Non-paretic* | 49 (75.4%) | N/A |
|  | *\*Unknown* | 1 (1.5%) | N/A |

\*data not available for one participant

### Self-reported Acceptance of Wearable Devices in Participants with Stroke

We show percentages of responses (Strongly Disagree, Disagree, Neutral, Agree, Strongly Agree) to each of the 31 TAQ statements rated by Likert scales organized by dimension (Figure 2). The five statements with the highest proportions of “Agree” or “Strongly Agree” responses were (in order; 2a, 2b, and so forth indicates that there were multiple statements with the same proportions of responses):

1. Statement 1: “I think that monitoring my activity and health 24 hours a day, 7 days a week, can be a good thing.”
2. Statement 7: “I was able to perform my daily tasks as usual while wearing the device.”
3. Statement 11: “The battery life of the device meets my expectations.”
4. Statement 18: “I was able to put the device on in a reasonable amount of time.”
5. Statement 20: “I am comfortable with my health data being shared with equipment manufacturers as long as it is shared anonymously.”

**Figure 2.**
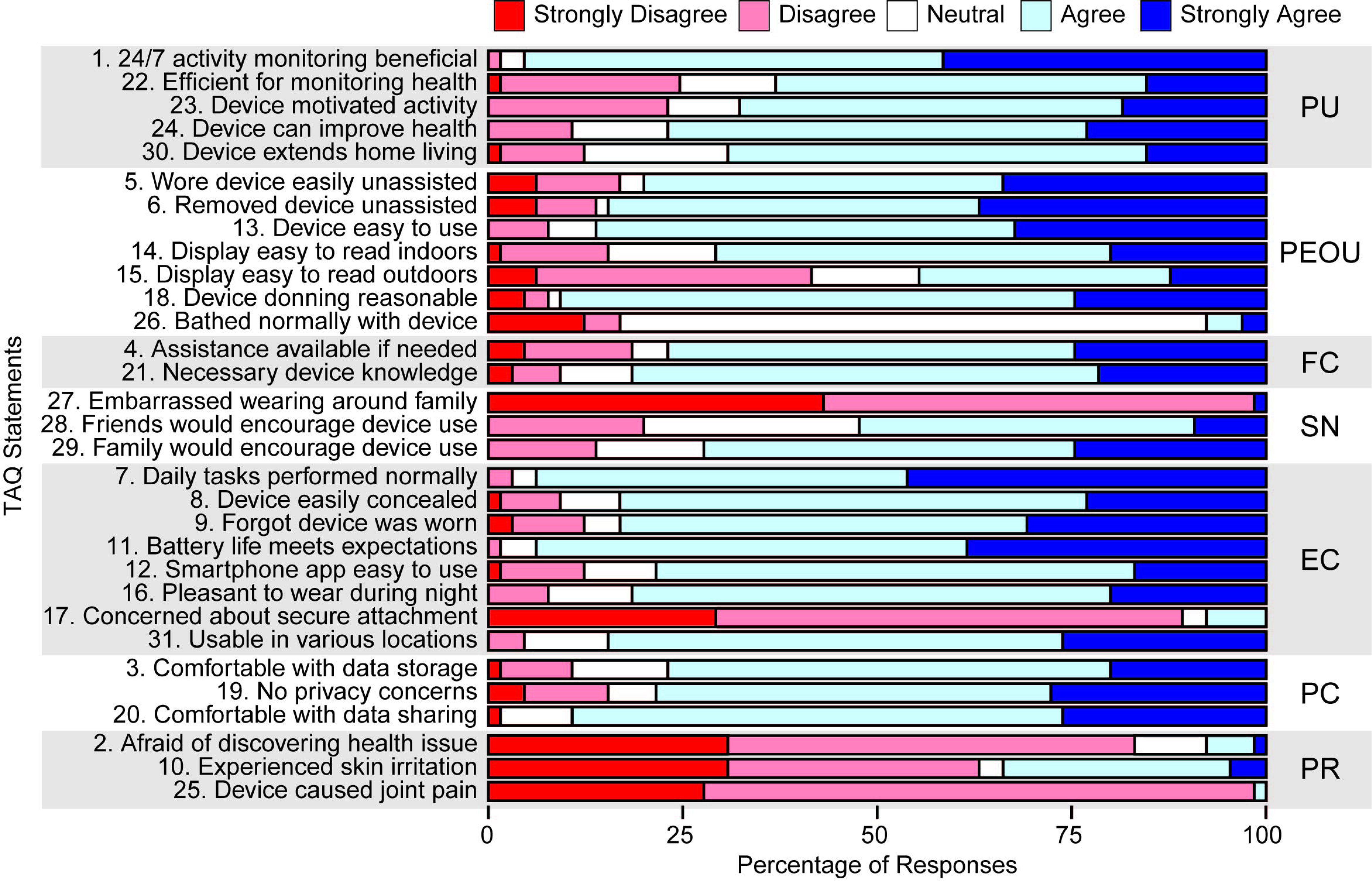
Percentages of responses to each individual statement on the TAQ grouped by the seven TAQ dimensions.

The five statements with the lowest proportions of “Agree” or “Strongly Agree” responses were (in order):

1. Statement 25: “Wearing the device caused me to have joint pain.”
2. Statement 27: “I was embarrassed to wear the device in front of family members.” 3a) Statement 2: “I was afraid that the device would discover a major health issue.” 3b) Statement 17: “I was concerned that the device is not securely attached to me.”
3. Statement 26: “I was able to shower or bathe normally while wearing the device.” As statements 1a-3b are reverse-coded; thus, a lower proportion of “Agree” or “Strongly Agree” responses to these statements indicates a more positive perception of the device. With the exceptions of the five statements immediately above as well as statements 10 (“I experience skin irritations while wearing the device”) and 15 (“I find the display of the device easy to read outdoors”), a majority of participants responded either “Agree” or “Strongly Agree” to each of the remaining 24 statements. We also note that responses to statement 26 were likely influenced by our instructions to the participants to remove the device while bathing or showering.

We show group means, standard deviations, medians, and interquartile ranges of these scores for each of the seven dimensions in Table 2 (alongside the maximum and minimum possible scores for each dimension) and for each individual TAQ statement in Table 3.

**Table 2.** Statistics of the responses to TAQ dimensions and the full TAQ.

|  | **Mean±SD** | **Median** | **IQR** | **Max possible** | **Min possible** |
| --- | --- | --- | --- | --- | --- |
| Perceived Usefulness | 19.1±3.3 | 20 | 4 | 25 | 5 |
| Perceived Ease of Use | 25.7±3.9 | 25 | 5 | 35 | 7 |
| Facilitating Conditions | 7.7±1.5 | 8 | 1 | 10 | 2 |
| Subjective Norm* | 11.6±1.9 | 12 | 3 | 15 | 3 |
| Equipment Characteristic* | 32.5±3.5 | 32 | 5 | 40 | 8 |
| Privacy Concern | 11.8±2.0 | 12 | 3 | 15 | 3 |
| Perceived Risk* | 11.9±2.0 | 12 | 3 | 15 | 3 |
| Technology Acceptance<br>Questionnaire | 120.4±12.8 | 118 | 14 | 155 | 31 |

\*These dimensions have reverse items in the subscore. Note that PR consists of all reversed questions, therefore the entire dimension is reversed.

**Table 3.** Statistics of the responses to each individual statement of the TAQ.

| Statement | Mean±SD | Median | IQR |
| --- | --- | --- | --- |
| 1) 24/7 activity monitoring beneficial | 4.4±0.6 | 4 | 1 |
| 2) Afraid of discovering health issue* | 4.1±0.9 | 4 | 1 |
| 3) Comfortable with data storage | 3.9±0.9 | 4 | 0 |
| 4) Assistance available if needed | 3.8±1.1 | 4 | 0 |
| 5) Wore device easily unassisted | 3.9±1.2 | 4 | 1 |
| 6) Removed device unassisted | 4.0±1.1 | 4 | 1 |
| 7) Daily tasks performed normally | 4.4±0.7 | 4 | 1 |
| 8) Device easily concealed | 4.0±0.9 | 4 | 0 |
| 9) Forgot device was worn | 4.0±1.0 | 4 | 1 |
| 10) Experienced skin irritation* | 3.6±1.3 | 4 | 3 |
| 11) Battery life meets expectations | 4.3±0.6 | 4 | 1 |
| 12) Smartphone app easy to use | 3.8±0.9 | 4 | 0 |
| 13) Device easy to use | 4.1±0.8 | 4 | 1 |
| 14) Display easy to read indoors | 3.7±1.0 | 4 | 1 |
| 15) Display easy to read outdoors | 3.1±1.2 | 3 | 2 |
| 16) Pleasant to wear during night | 3.9±0.8 | 4 | 0 |
| 17) Concerned about secure attachment* | 4.1±0.8 | 4 | 1 |
| 18) Device donning reasonable | 4.0±0.9 | 4 | 0 |
| 19) No privacy concerns | 3.9±1.1 | 4 | 1 |
| 20) Comfortable with data sharing | 4.1±0.7 | 4 | 1 |
| 21) Necessary device knowledge | 3.9±0.9 | 4 | 0 |
| 22) Efficient for monitoring health | 3.5±1.1 | 4 | 1 |
| 23) Device motivated activity | 3.6±1.0 | 4 | 1 |
| 24) Device can improve health | 3.9±0.9 | 4 | 0 |
| 25) Device caused joint pain* | 4.3±0.5 | 4 | 1 |
| 26) Bathed normally with device | 2.8±0.8 | 3 | 0 |
| 27) Embarrassed wearing around family* | 4.4±0.7 | 4 | 1 |
| 28) Friends would encourage device use | 3.4±0.9 | 4 | 1 |
| 29) Family would encourage device use | 3.8±1.0 | 4 | 1 |
| 30) Device extends home living | 3.7±0.9 | 4 | 1 |
| 31) Usable in various locations | 4.0±0.8 | 4 | 1 |

\*These statements have been reverse coded to preserve directionality.

Medians of the summed scores for each dimension ranged from 71% in PEOU (median=25, maximum possible score=35) to 80% in all other dimensions (Table 2). Furthermore, all dimensions – with the exception of PEOU – showed median scores of 4 (or “Agree”) on each individual statement (Table 3). Modestly lower scores in the PEOU dimension were largely driven by generally less agreeable responses to statements 15 “I find the display of the device easy to read outdoors” and 26 “I was able to shower or bathe normally while wearing the device”, again noting that we instructed the participants to remove the device before bathing or showering. The median for the total TAQ score was 76% of the maximum possible score (median=118, maximum possible score=155).

Next, we analyzed responses to the six multiple choice questions from the TAQ (questions 32-37) that did not have designated dimensions (Table 4). Most participants found the device useful (question 32), with 95.4% rating the information provided as either “very useful” or “somewhat useful”. Nearly all participants (92.3%) expressed willingness to continue using the device to monitor their health (question 33), and 96.9% reported wearing the device for 15–21 days out of the 21-day period (question 34). In terms of value, most participants indicated willingness to pay no more than $100 for the device (question 35). Most (90.8%) participants reported looking at their health data provided by their device (question 36). Finally, self-perception of activity levels varied among participants with 64.6% considering themselves to be active (question 37).

**Table 4.**
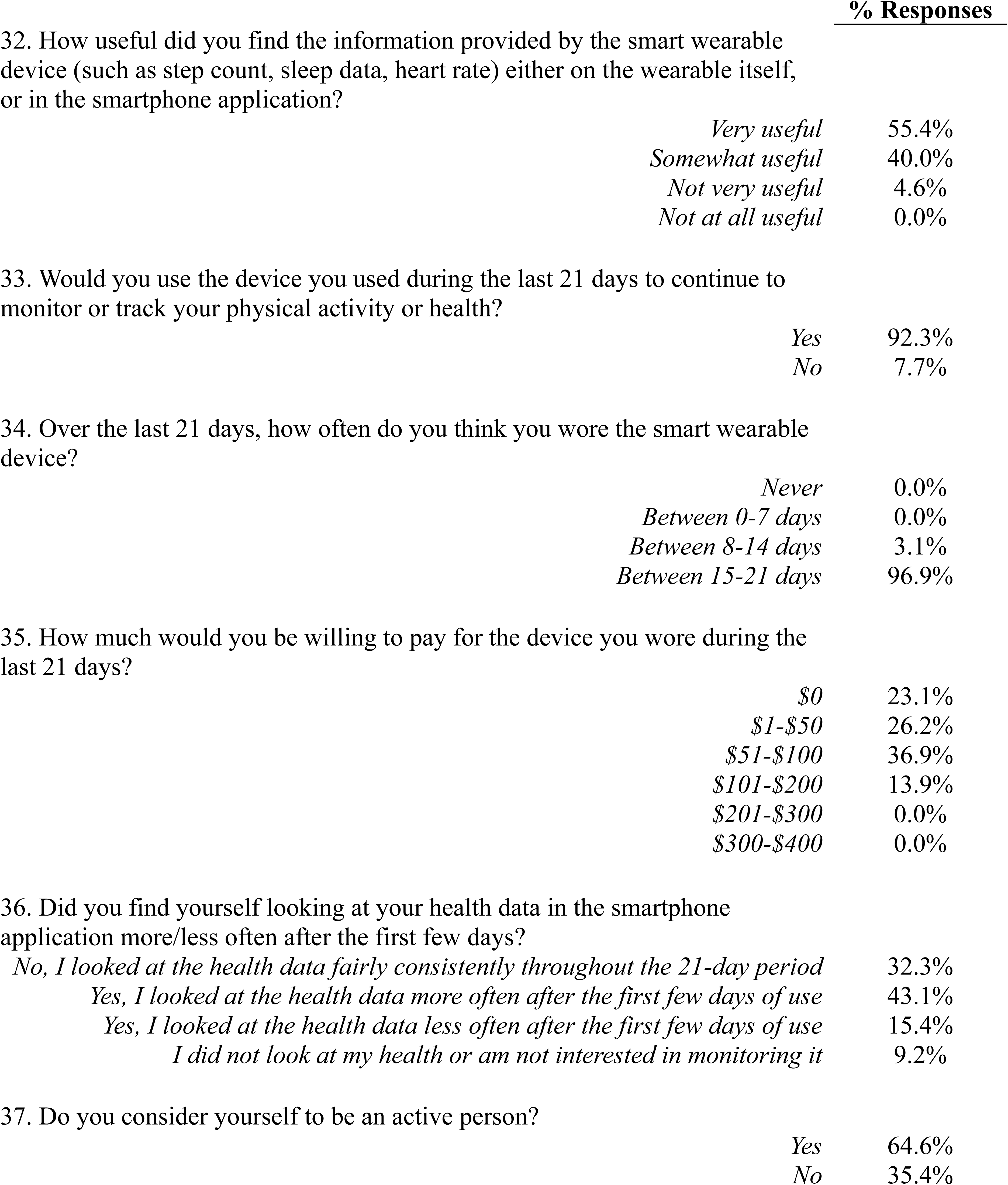
Responses to TAQ multiple choice questions.

### Relationships Between Self-Reported Acceptance and Fitbit Adherence

Overall participants wore the Fitbit an average and standard deviation of 80.0%±24.7% of the total minutes in a day, with a median wear time of 91% and an interquartile range of 22%. Wear time exceeded the threshold needed to be considered a valid wear day on 78.0%±25.8% of days with a median of 90% and an interquartile range of 33%. The scatterplots in Figures 3 and 4 show relationships between the summed scores of the different TAQ dimensions (as well as TAQ total scores) and the percentages of wear time and valid wear days, respectively. Contrary to our hypothesis, there were no statistically significant positive relationships between TAQ dimension summed scores or total scores and Fitbit adherence metrics. We did, however, observe two small but statistically significant negative associations between PU and percent wear time (ρ=−0.27, p=0.03) and between PU and the percentage of valid wear days (ρ=−0.26, p=0.04).

**Figure 3.**
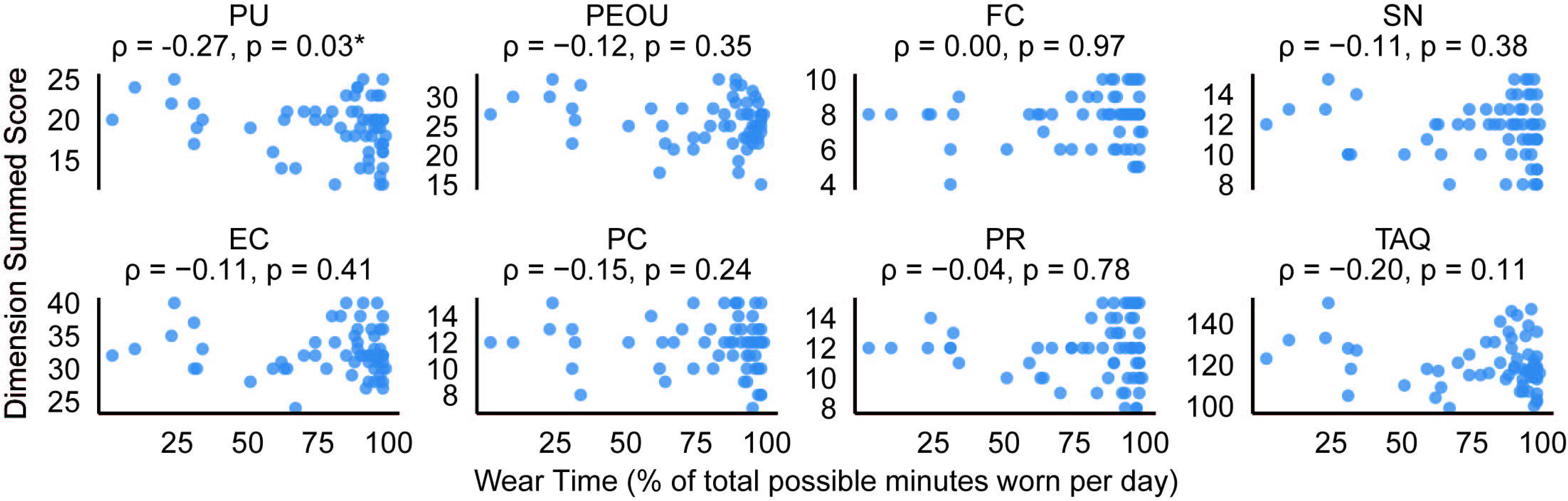
Scatterplots showing relationships between summed scores for each TAQ dimension (as well as total TAQ scores) and Fitbit wear time. Asterisks indicate statistically significant relationships (p<0.05).

**Figure 4.**
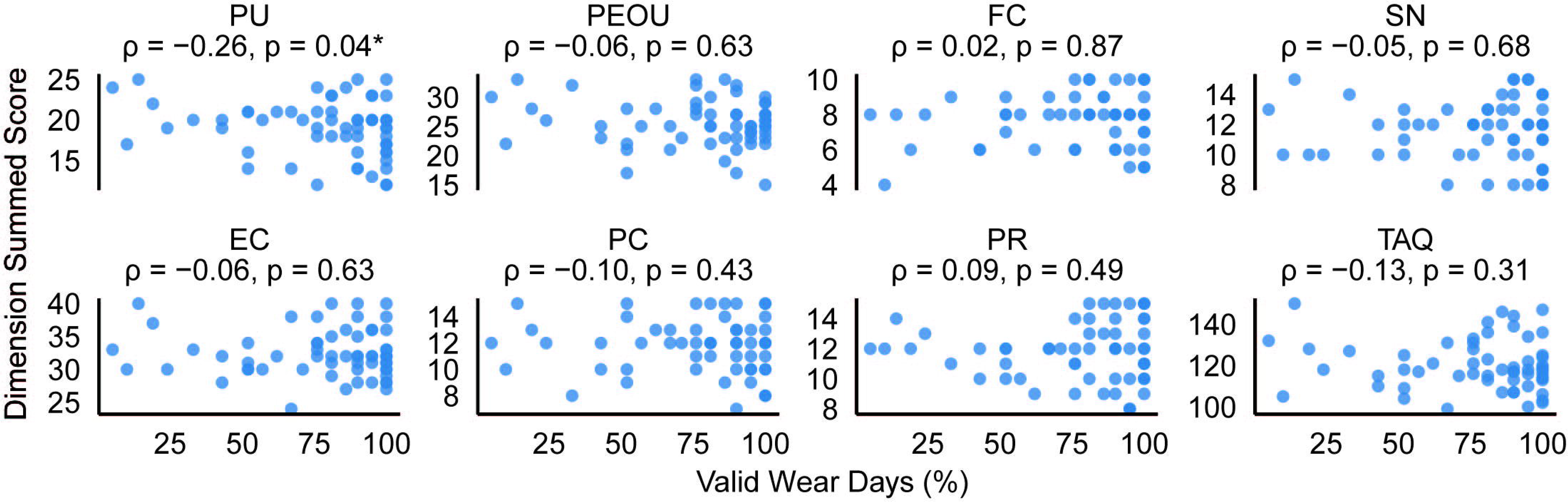
Scatterplots showing relationships between summed scores for each TAQ dimension (as well as total TAQ scores) and valid Fitbit wear days. Asterisks indicate statistically significant relationships (p<0.05).

## Discussion

### Principal Results

Our study examines self-reported perceptions of the Fitbit Inspire 2 wearable activity monitor among individuals with stroke, as measured by the Technology Acceptance Questionnaire (TAQ). A majority of the participants thought the device was beneficial for their health, efficient for monitoring their health, easy to use and don/doff, and unintrusive to daily life; one notable exception was the response to the statement “I find the display of the device easy to read outdoors”. Generally, participants did not express significant concerns about privacy or data security, consistent with prior studies.[30,31] Contrary to our hypothesis, more agreeable responses to the TAQ statements were not associated with higher device percentage of average daily wear time and percentage of valid wear days at a statistically significant level.

### Comparison with Prior Work

The findings of our study are consistent with previous literature demonstrating acceptance of wearable activity monitors in other populations.[13,32–36] Given the growing interest in using wearables for activity monitoring and telerehabilitation after stroke,[26,37–40] it is important to consider not only their potential benefits but also because of low levels of physical activity, cognitive and motor impairments that may affect this population’s acceptance and engagement.[10,41,42] There were no commonly reported acceptance concerns that we deemed likely to be related to post-stroke motor impairment: as examples, participants widely agreed to the statements “I was able to wear the device easily without help from another person,” “I find the device easy to use,” “I was able to put the device on in a reasonable amount of time,” and “I was able to remove the device easily without help from another person.” These findings complement recent studies demonstrating the perceived value and user satisfaction of wearable technologies in stroke rehabilitation[43–45] and support a path toward scalable implementation of remote monitoring with wearable devices. This is likely due in part to the flexibility we allowed in permitting participants to wear the device on their paretic side if needed, accommodating individual motor abilities.

We also highlight that the study participants reported generally agreeable responses across all seven dimensions of the TAQ. Previous work highlighted that technical and usability issues (e.g., requiring a mobile application to sync the data from the device to the server) may affect the perceived usefulness of wearable devices;[16] however, we did not observe this in our sample. This is potentially because any technical difficulties were often addressed via interactions with the study team. Our findings across the different dimensions were largely similar to those reported in a previous sample of older adults.[13] It is important to emphasize that monitoring of device adherence may be necessary despite the high reported acceptance. Our findings did not support the hypothesis that higher user acceptance as measured by the TAQ would correlate with measures of adherence to wearing the device, as we did not observe statistically significant positive correlations between TAQ responses and our measures of adherence. This revealed that high reported acceptance of the device does not guarantee that a patient or research participant will necessarily adhere to wearing the device in everyday life. Technologies that help to automate oversight of device wear and messaging to promote adherence will be important for ensuring data quality.[46]

The correlational analyses indicated small but significant negative associations between the perceived usefulness (PU) score and both adherence metrics. These results are contrary to our hypotheses, which were grounded in the TAM and related literature, where PU is typically positively associated with adoption behaviors.[47,48] Existing studies have shown that higher PU is often associated with sustained use of technology across various domains,[49–51] including healthcare settings.[52–55] Our findings may be attributed to the specific context in which the wearable device was used. Unlike most prior technology adoption studies where participants voluntarily adopted technology based on its usefulness, our study cohort used the Fitbit as part of their participation in a research study. This mandated context could have influenced the PU from typical motivational factors driving technology adoption. It is possible that participants did not view Fitbit as inherently useful for their health recovery goals but instead perceived it as a tool for fulfilling study requirements. While survey responses suggest that most participants agreed that the device could improve health and monitor well-being efficiently, these endorsements may reflect a general perception of health technology utility rather than a personalized after stroke recovery needs. Consequently, their assessment of PU may reflect this externally driven motivation rather than genuine alignment with personal health management goals.

As the push toward clinical use of wearables in stroke rehabilitation continues to move forward, it is also important to consider the needs and perspectives of all key stakeholders – patients, their family members and caregivers, and clinicians – in addition to the device acceptance reported by patients in our study. For example, recent studies have provided vital information regarding how persons with stroke prefer to receive data from wearables and identified a set of metrics deemed most useful;[23] incorporated perspectives from patients and clinicians on the value of using wearables and identified preferences for incorporation into clinical care;[44] and demonstrated important design considerations for adoption of the wearables and accompanying smartphone applications as outlined by persons with stroke.[45] For example, to consider the difficulty of donning/doffing providing an alternative strap such as a Velcro strap instead of the original buckle band. Future work should consider these multifaceted aspects – patient acceptance, patient and clinician data provision preferences, and device design – as we move closer to clinical implementation of wearables in stroke rehabilitation.

### Limitations

We acknowledge some limitations in our study. First, our sample was heterogeneous regarding stroke chronicity. Accordingly, we did not design this study to assess how wearable device acceptance may differ across stages of stroke recovery (e.g., acute, subacute, chronic). However, this diversity in stroke recovery stages could be beneficial, as it reflects the clinical reality where wearable devices in stroke rehabilitation should not discriminate based on recovery stage but rather be accessible and valuable to patients at various points in their recovery. Second, we only used the Fitbit Inspire 2 device. While we anticipate that many of our findings may generalize across different models of commercially available wearable devices due to the nature of the statements included in the TAQ, we do not have data to support this directly. Third, we focused on the TAQ to provide information about device acceptance in particular. We did not use other instruments that could provide additional data on other aspects of patient perceptions about wearable devices (e.g., the System Usability Scale for assessment of usability).

## Conclusion

This study reported the perceived acceptance of a wrist-worn activity monitor in persons with stroke. In response to statements on the TAQ, participants with stroke generally reported the device to be beneficial for their health, useful for monitoring their health, easy to use, and minimally intrusive. We observed generally agreeable responses to TAQ statements across all seven dimensions of the instrument. Contrary to our hypothesis, more agreeable responses to the TAQ statements were not positively correlated with metrics of device wear, revealing that adherence to wearing the device should not be assumed even when participants report high device acceptance. In summary, this study detailed new information about the acceptance of wearable activity monitors in persons with stroke and its association with real-world device wear.

## Data Availability

All data produced in the present study are available upon reasonable request to the authors

## Acknowledgments

We acknowledge funding from the American Heart Association (grants 23IPA1054140 and 935556 to RTR, grant 24POST1187285 to GCB) and the Sheikh Khalifa Stroke Institute at Johns Hopkins Medicine.

## Abbreviations

CONSORT: Consolidated standards of reporting trials
EC: Equipment characteristics
FC: Facilitating conditions
IQR: Interquartile range
PC: Privacy concern
PEOU: Perceived ease of use
PR: Perceived risk
PU: Perceived usefulness
SN: Subjective norm
TAM: Technology Acceptance Model
TAQ: Technology Acceptance Questionnaire

