## Supplementary material for "Self-reported acceptance of a wearable activity monitor in persons with stroke": Descriptive on additional TAQ questions (from another version of this preprint: version 2, 2025-05-03)

**Table 4. Responses to TAQ multiple choice questions.**

|  | **% Responses** |
| --- | --- |
| 32. How useful did you find the information provided by the smart wearable device (such as step count, sleep data, heart rate) either on the wearable itself, or in the smartphone application? |  |
| *Very useful* | 55.4% |
| *Somewhat useful* | 40.0% |
| *Not very useful* | 4.6% |
| *Not at all useful* | 0.0% |
| 33. Would you use the device you used during the last 21 days to continue to monitor or track your physical activity or health? |  |
| *Yes* | 92.3% |
| *No* | 7.7% |
| 34. Over the last 21 days, how often do you think you wore the smart wearable device? |  |
| *Never* | 0.0% |
| *Between 0-7 days* | 0.0% |
| *Between 8-14 days* | 3.1% |
| *Between 15-21 days* | 96.9% |
| 35. How much would you be willing to pay for the device you wore during the last 21 days? |  |
| *$0* | 23.1% |
| *$1-$50* | 26.2% |
| *$51-$100* | 36.9% |
| *$101-$200* | 13.9% |
| *$201-$300* | 0.0% |
| *$300-$400* | 0.0% |
| 36. Did you find yourself looking at your health data in the smartphone application more/less often after the first few days? |  |
| *No, I looked at the health data fairly consistently throughout the 21-day period* | 32.3% |
| *Yes, I looked at the health data more often after the first few days of use* | 43.1% |
| *Yes, I looked at the health data less often after the first few days of use* | 15.4% |
| *I did not look at my health or am not interested in monitoring it* | 9.2% |
| 37. Do you consider yourself to be an active person? |  |
| *Yes* | 64.6% |
| *No* | 35.4% |
